# Q3: An ICU Acuity Score for Exploring the Effects of Changing Acuity Throughout the Stay

**DOI:** 10.1101/2022.03.05.22271962

**Authors:** Stephen E. Brossette, Ning Zheng, Daisy Y. Wong, Patrick A. Hymel

## Abstract

**Intro:** We develop a straightforward ICU acuity score (Q3) that is calculated every 3 hours throughout the first 10 days of the ICU stay. Q3 uses components of the Oxford Acute Severity of Illness Score (OASIS) and incorporates a new component score for vasopressor use. In well-behaved models of ICU mortality, the marginal effects of Q3 are significant across the first 10 days of the ICU stay. In separate models, Q3 has significant effects on ICU remaining length of stay. The score has implications for work that seeks to explain modifiable mechanisms of changing acuity during the ICU stay.

**Methods:** From the MIMIC-III database, select ICU stays from 5 adult ICUs were partitioned into consecutive 3-hour segments. For each segment, the number of vasopressors administered and all 10 OASIS component scores were computed. Models of ICU mortality were estimated. OASIS component effects were examined, and vasopressor count bins were weighted. Q3 was defined as the sum of 8 retained OASIS components and a new weighted vasopressor score. Models of ICU mortality quadratic in Q3 were estimated for each of the first 10 ICU days and were subjected to segment-level, location-specific tests of discrimination and calibration on newer ICU stays. Marginal effects of Q3 were computed at different levels of Q3 by ICU day, and average marginal effects of Q3 were computed at each location by ICU day. ICU remaining length of stay (LOS) models were also estimated and the effects of Q3 were similarly examined.

**Results:** Daily ICU mortality models using Q3 show no evidence of misspecification (Pearson-Windmeijer p>0.05, Stukel p>0.05), discriminate well in all ICUs over the first 10 days (AUROC ∼ 0.72 – 0.85), and are generally well calibrated (Hosemer-Lemeshow p>0.05, Spiegelhalter’s z p>0.05). A one-unit increase in Q3 from typical levels (Q3=15) affects the odds of ICU mortality by a factor of 1.14 to 1.20, depending on ICU day (p<0.001), and the ICU remaining LOS by 5.8 to 9.6% (p<0.001). On average, a one-unit increase in Q3 increases the probability of ICU mortality by 1 to 2 percentage points depending on location and ICU day, and ICU remaining LOS by 5 to 10 hours depending on location and ICU day.

**Conclusion:** Q3 significantly affects ICU mortality and ICU remaining LOS in different ICUs across the first 10 days of the ICU stay. Depending on location and ICU day, a one-unit increase in Q3 increases the probability of ICU mortality by or 1-2 percentage points and ICU remaining LOS by 5 to 10 hours. Unlike static acuity scores or those updated infrequently, Q3 could be used in explanatory models to help elucidate mechanisms of changing ICU acuity.

## Introduction

### Background and Significance

Many ICU acuity scores have been developed [1-7]. They equate acuity with odds of mortality (in-ICU or in-hospital) and are used in models that discriminate well and are sometimes well calibrated. Acuity scores differ in variables used, but most use vital signs, laboratory results, medications, ventilator status, comorbidities, and patient demographics. Established ICU acuity scores include APACHE, SAPS, MPM, SOFA, and OASIS, amongst others. Performance is typically evaluated at the end of the first ICU day (except SOFA which is also evaluated every subsequent 48 hours) with discrimination evaluated by AUROC/c-statistics, and calibration examined via Hosemer-Lemeshow (HL) tests.

The Oxford Acute Severity of Illness Score (OASIS) is a relatively new ICU acuity score. It uses less data than others (10 variables), does not depend on laboratory results which can lag in acquisition and reporting, and performs about as well as other acuity scores [7]. It was designed to be more computable and timelier than other scores, but like most other scores, has only been evaluated at the end of the first ICU day.

Real-time models of ICU acuity have also been developed [8-10]. They typically use many clinical variables instead of a single summarized acuity score, and rely on machine learning techniques to capture complex, non-linear relationships between explanatory variables and the outcome. While they typically perform well, machine learning models are (so far) uninterpretable, and the marginal effects of key variables on the odds of mortality are complex and unknown. It is also notable that in a study by Johnson and Mark [10], machine learning techniques only slightly outperformed logistic regression models, which are interpretable.

In this study we construct a new ICU acuity score (Q3) that uses a subset of OASIS component scores and incorporates a new component score based on vasopressor use. Q3 is computed every 3 hours throughout the ICU stay. We show that straightforward models quadratic in Q3 are well-behaved over the first 10 ICU days across different ICUs, and that the marginal effects of Q3 on ICU mortality and ICU remaining LOS are significant and relatively stable.

## Materials and Methods

### Data

We analyzed data from the MIMIC-III database, version 1.4. MIMIC-III is comprised of de-identified data from over forty thousand patients who stayed in critical care units of the Beth Israel Deaconess Medical Center between 2001 and 2012 [11-12]. It contains data from Carevue and Metavision ICU systems, the laboratory information system, and the hospital EHR. Carevue data are from years 2001-2008 and Metavision data are from years 2008-2012, with very little overlap.

We used adult (>=18yo) ICU stays from the MICU (medical), SICU (surgical), CCU (coronary care), CSRU (cardiac surgery recovery), and TSICU (trauma surgical) (n=52,061). Each stay was divided into contiguous, non-overlapping 3-hour *segments*, indexed from 0.

### Computing Q3 component scores

For each 3-hour segment, we computed the number of distinct vasopressors administered along with all 10 OASIS component scores. The Oxford Acute Severity of Illness Score (OASIS) is the sum of the first-day maximum scores of 10 component scores, described by Johnson, et. al [7]. Using the same component bins and weights (Fig 1), we computed each component score at the end of each segment for 80 segments (10 days) using the latest-available data, without considering relative score values over time. Specifically, each component score used the latest data over the previous 24 hours, except for *preiculos, age*, and *electivesurg*, which were constant throughout the stay. If no component data existed within 24 hours, the component score was set to a default value of 0. For all components except *urineout*, the implementation of the latest-data logic was straightforward. For *urineout*, the 24-hour rate of urine output, the following logic was used. At each urine output event, a 24-hour rate of urine output was computed by dividing the volume of urine collected since the last event by the number of elapsed hours since that event or the start of the ICU stay, if no event existed. This rate was assigned to the time window from the current output event back to the previous event (or start of the ICU stay). Then for each segment, *urineout* was computed as the average of all 24-hour rates assigned to time windows that overlapped with the segment. If the end of a segment occurred after the last urine output event, *urineout* was set the prior value of *urineout*.

**Figure 1:**
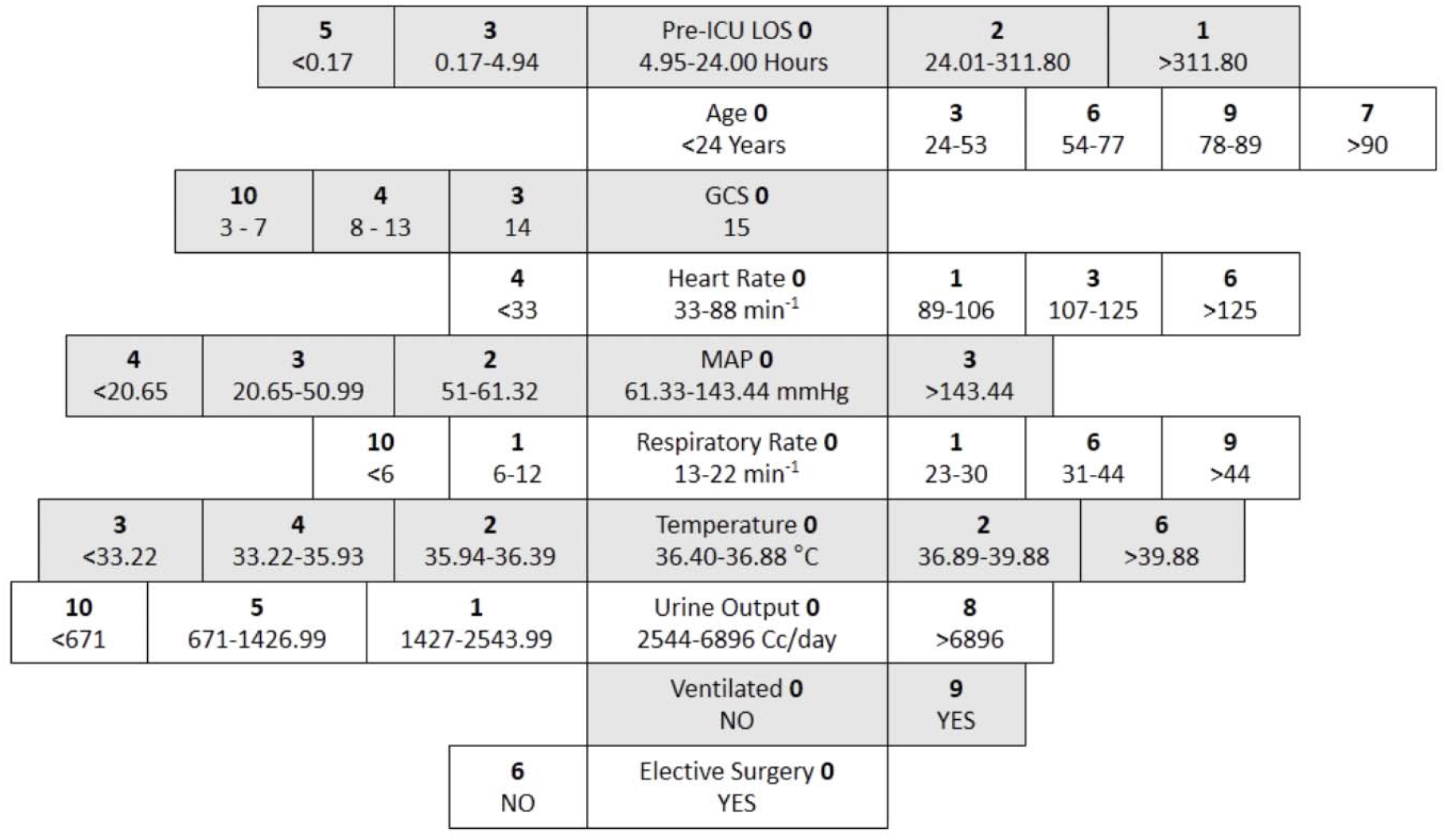
Component bins and weights for the Oxford Acute Severity of Illness Score (OASIS) [7]

Vasopressors were comprised of norepinephrine, epinephrine, vasopressin, dobutamine, dopamine, milrinone, and phenylephrine. For each segment, the number of distinct vasopressors used was binned into none, one, two, and 3 or more (Table 1).

**Table 1:** Vasopressor use in first 10 ICU days

| n<br>vasopressors | % segments |
| --- | --- |
| 0 | 82.2 |
| 1 | 13.6 |
| 2 | 3.2 |
| ≥3 | 1.1 |

### Selecting Q3 components and weighting vasopressor use bins

The effects of OASIS components and vasopressor use on ICU mortality were examined through estimates of Equation A.

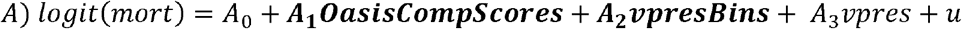

In this equation, *logit* is the logistic function; *mort* is an indicator for ICU mortality; ***OasisCompScores*** is a vector of the 10 OASIS component scores computed at each segment; ***vpresBins*** is a vector of 4 indicators for the number of distinct vasopressors administered in the segment (n=0,1,2,>=3); and *vpres* is the weight assigned to the vasopressor bin for the number of vasopressors administered in the segment. The four vasopressor bins were weighted by first estimating Equation A without *vpres* and using the parameter estimates of ***vpresBins*** to calculate a starting weight for each bin. Then a simple integer-grid-search around the starting weights was executed until estimates of Equation A (with *vpres*) produced *vpres* effects comparable to those of other significant components and ***vpresBins*** effects that were insignificant. Throughout, Equation A was estimated once for each of the first 10 ICU days, using only the fourth segment of each day to avoid repeated sampling of the same patients. Parameter estimates across all 10 days were examined and a subset of components with consistent and significant effects was retained. Finally, Q3 was defined as the sum of the retained components.

### Estimating the effects of Q3 on ICU mortality

The effects of Q3 on ICU mortality were examined through estimates of Equation B.

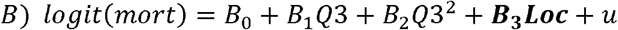

The new variable ***Loc*** is a vector of 5 location indicators for MICU, SICU, CCU, CSRU, and TSICU. Equation *B* was estimated 10 times using Carevue stays (2001-2008), once for each of the first 10 ICU days, using data from the fourth segment of each day. Pearson-Windmeijer [13] and Stukel [14] goodness-of-fit tests were used to check for sources of misspecification.

### Discrimination and calibration testing

Discrimination is a measure of a model’s ability to assign higher scores to patients with events (e.g. death) than without, and calibration is a measure of a model’s ability to accurately estimate the number of events in different quantiles of probability. The 10 estimated daily models of Equation *B* (using Carevue stays, 2001-2008) were tested for discrimination and calibration on strictly newer Metavision stays (2008-2012). By ICU, the day 0 model was tested at each segment 0 through 7, the day 1 model at segments 8 through 15, etc. Discrimination was evaluated by AUROC/c-statistics and calibration by Hosemer-Lemeshow (HL) tests using 10 groups. Since HL tests become too powerful in large samples [15], some have recommended that HL tests use smaller random samples where appropriate [15-18]. Although exact sample size recommendations are unavailable, and the determination of calibration is ultimately subjective, one strategy suggested by Paul et. al. is to perform HL tests on random samples of size n=1000 and 10 groups [16], and others have employed random sampling for HL calibration tests [17-18]. We follow this strategy and use a random sample of n=1000 observations when more than 1000 observation were available. An insignificant HL test (p>0.05) is suggestive of sufficient segment-level calibration. Overall model calibration was deemed sufficient if, in a single ICU, segment-level calibration results were mostly insignificant over extended periods of time. In addition to c-statistics and HL tests, Brier scores were computed for each segment and location. Brier scores simultaneously capture features of discrimination and calibration [19]. The Spiegelhalter’s z-test was used to test for Brier score significance, and p-values were computed.

### Marginal and average marginal effects

Since Equation B is quadratic in Q3, the marginal effects of Q3 depend on the level of Q3 itself as shown by the derivative of Equation B with respect to Q3: d(logit(*mort*))/dQ3 = B_1_ + 2B_2_Q3. Marginal effects of the 10 daily models were calculated at Q3=5, 15, 25, and 35. The average marginal effects of Q3 (AME, in probability points) were calculated daily.

### Estimating the effects of Q3 on ICU remaining LOS

The effects of Q3 on ICU remaining LOS (rLOS) were examined through Poisson regression where the distributional parameter lambda is given by the following equation.

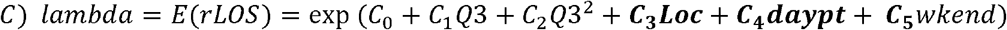

Here, *rLOS* is the remaining LOS (in hours) from the end of the current segment, ***daypart*** is a vector of indicators for 6 consecutive 4-hour intervals (starting from midnight) in which the segment starts, and *wkend* is an indicator for the segment starting on Saturday or Sunday. Equation *C* was estimated once daily for each of the first 10 ICU days. Robust standard errors were employed to adjust for any overdispersion and are valid under *any* conditional variance assumption [20]. The natural log of equation C is quadratic in Q3 and its marginal effects depend on the level of Q3. They were estimated at Q3=5, 15, 25, and 35 and can be easily interpreted as proportional changes in rLOS. Average marginal effects (in hours) for each ICU day and location were also computed.

All analysis was done using Stata v16 (StataCorp. 2019. *Stata Statistical Software: Release 16*. College Station, TX: StataCorp LLC). Investigators were certified to use the MIMIC-III database. All data were previously de-identified, and the public use of MIMIC has been approved by the IRB of its hosting organization. No additional IRB approval was required. Project code is available from the authors.

## Results

Selected ICU stays are summarized in Table 2. ICU mortality rates differ between ICUs and are higher in Carevue stays. Seventy-fifth percentile ICU LOS also differ by location and are higher in Carevue stays. Other descriptive statistics of the MIMIC III data can be found in Dai et al [21].

**Table 2:** Selected ICU stays (Carevue 2001-2008; Metavision 2008-2012)

|  | n | ICU LOS [IQR] (d) | ICU Mortality (%) |
| --- | --- | --- | --- |
| All | (28,817; 23,244) | ([0.75, 4.5]; [1.0, 3.75]) | (9.1; 7.7) |
| MICU | (10,399; 10,090) | ([1.1, 4.5]; [1.0, 3.5]) | (11.6; 9.0) |
| SICU | (4,418; 4,277) | ([1.3, 5.5]; [1.0, 4.1]) | (10.1; 8.3) |
| CCU | (4,778; 2,731) | ([1.1, 4.1]; [1.1, 3.9]) | (9.3; 8.7) |
| CSRU | (5,799; 3,346) | ([1.1, 4.0]; [1.1, 3.1]) | (3.5; 2.7) |
| TSICU | (3,423; 2,800) | ([1.0, 4.9]; [1.1; 4.1]) | (9.6; 7.0) |

For each observation, *vpres* was assigned the value (weight) that corresponds to the number of distinct vasopressors administered in the segment (Table 3). From Equation A, the estimated marginal effects (as odds ratios, OR) of ***OasisCompScores, vpresBins***, and *vpres* by ICU day are shown in Table 4.

**Table 3:** Vasopressor bins and weights

| n vasopressors<br>(bins) | *vpres*<br>(weights) |
| --- | --- |
| 0 | 0 |
| 1 | 4 |
| 2 | 8 |
| >=3 | 10 |

**Table 4:**
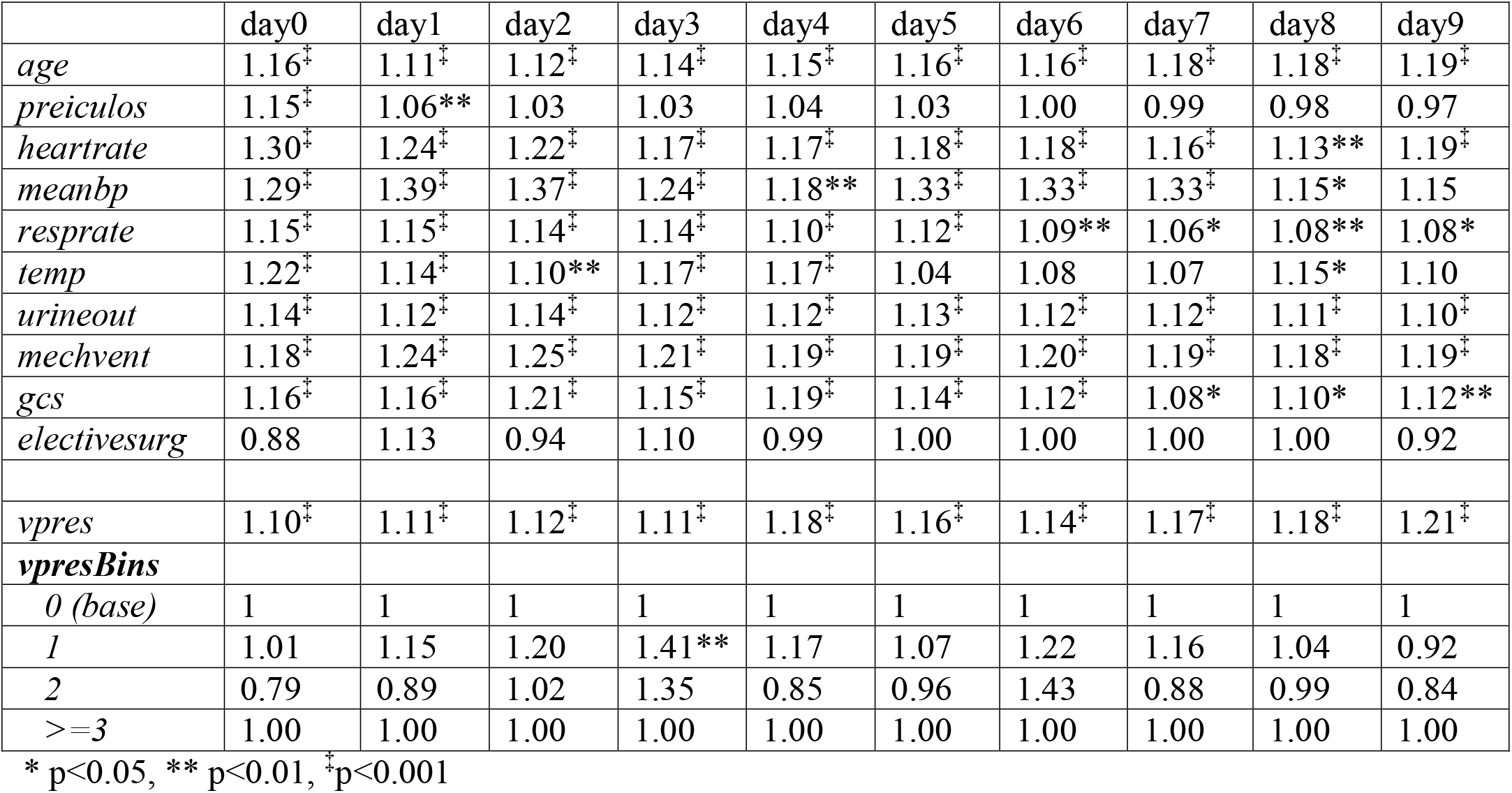
Estimated effects (OR) of component scores on odds of ICU mortality (Eq A)

|  | day0 | day1 | day2 | day3 | day4 | day5 | day6 | day7 | day8 | day9 |
| --- | --- | --- | --- | --- | --- | --- | --- | --- | --- | --- |
| *age* | 1.16<sup>‡</sup> | 1.11<sup>‡</sup> | 1.12<sup>‡</sup> | 1.14<sup>‡</sup> | 1.15<sup>‡</sup> | 1.16<sup>‡</sup> | 1.16<sup>‡</sup> | 1.18<sup>‡</sup> | 1.18<sup>‡</sup> | 1.19<sup>‡</sup> |
| *preiculos* | 1.15<sup>‡</sup> | 1.06** | 1.03 | 1.03 | 1.04 | 1.03 | 1.00 | 0.99 | 0.98 | 0.97 |
| *heartrate* | 1.30<sup>‡</sup> | 1.24<sup>‡</sup> | 1.22<sup>‡</sup> | 1.17<sup>‡</sup> | 1.17<sup>‡</sup> | 1.18<sup>‡</sup> | 1.18<sup>‡</sup> | 1.16<sup>‡</sup> | 1.13** | 1.19<sup>‡</sup> |
| *meanbp* | 1.29<sup>‡</sup> | 1.39<sup>‡</sup> | 1.37<sup>‡</sup> | 1.24<sup>‡</sup> | 1.18** | 1.33<sup>‡</sup> | 1.33<sup>‡</sup> | 1.33<sup>‡</sup> | 1.15* | 1.15 |
| *resprate* | 1.15<sup>‡</sup> | 1.15<sup>‡</sup> | 1.14<sup>‡</sup> | 1.14<sup>‡</sup> | 1.10<sup>‡</sup> | 1.12<sup>‡</sup> | 1.09** | 1.06* | 1.08** | 1.08* |
| *temp* | 1.22<sup>‡</sup> | 1.14<sup>‡</sup> | 1.10** | 1.17<sup>‡</sup> | 1.17<sup>‡</sup> | 1.04 | 1.08 | 1.07 | 1.15* | 1.10 |
| *urineout* | 1.14<sup>‡</sup> | 1.12<sup>‡</sup> | 1.14<sup>‡</sup> | 1.12<sup>‡</sup> | 1.12<sup>‡</sup> | 1.13<sup>‡</sup> | 1.12<sup>‡</sup> | 1.12<sup>‡</sup> | 1.11<sup>‡</sup> | 1.10<sup>‡</sup> |
| *mechvent* | 1.18<sup>‡</sup> | 1.24<sup>‡</sup> | 1.25<sup>‡</sup> | 1.21<sup>‡</sup> | 1.19<sup>‡</sup> | 1.19<sup>‡</sup> | 1.20<sup>‡</sup> | 1.19<sup>‡</sup> | 1.18<sup>‡</sup> | 1.19<sup>‡</sup> |
| *gcs* | 1.16<sup>‡</sup> | 1.16<sup>‡</sup> | 1.21<sup>‡</sup> | 1.15<sup>‡</sup> | 1.19<sup>‡</sup> | 1.14<sup>‡</sup> | 1.12<sup>‡</sup> | 1.08* | 1.10* | 1.12** |
| *electivesurg* | 0.88 | 1.13 | 0.94 | 1.10 | 0.99 | 1.00 | 1.00 | 1.00 | 1.00 | 0.92 |
| *vpres* | 1.10<sup>‡</sup> | 1.11<sup>‡</sup> | 1.12<sup>‡</sup> | 1.11<sup>‡</sup> | 1.18<sup>‡</sup> | 1.16<sup>‡</sup> | 1.14<sup>‡</sup> | 1.17<sup>‡</sup> | 1.18<sup>‡</sup> | 1.21<sup>‡</sup> |
| ***vpresBins*** |  |  |  |  |  |  |  |  |  |  |
| *0 (base)* | 1 | 1 | 1 | 1 | 1 | 1 | 1 | 1 | 1 | 1 |
| *1* | 1.01 | 1.15 | 1.20 | 1.41** | 1.17 | 1.07 | 1.22 | 1.16 | 1.04 | 0.92 |
| *2* | 0.79 | 0.89 | 1.02 | 1.35 | 0.85 | 0.96 | 1.43 | 0.88 | 0.99 | 0.84 |
| *>=3* | 1.00 | 1.00 | 1.00 | 1.00 | 1.00 | 1.00 | 1.00 | 1.00 | 1.00 | 1.00 |

\* p<0.05, \*\* p<0.01, <sup>‡</sup>p<0.001

With the vasopressor bin weights in Table 3, and *vpres* set to the weight of its corresponding bin, *vpres* effects are approximately the right size, as designed. At the same time, ***vpresBins*** indicators are mostly insignificant, also as designed, suggesting that no additional information on vasopressor use remains in the ***vpresBins*** indicators. The *vpres* component score is very significant as expected since vasopressors are important in critical care medicine. OASIS component scores for *heartrate, meanbp, resprate, temp, urineout, mechvent, gcs, and age* are frequently significant and are in the expected direction (OR>1). *PreICUlos* is significant only on the first two ICU days. *Electivesurgery* is not significant.

Q3 was defined as the sum of 9 component scores: *vpres, heartrate, meanbp, resprate, temp, urineout, mechvent, gcs, and age. PreICUlos* and *Electivesurgery* were not used. Boxplots of Q3 by ICU across ICU days are shown in Fig 2. Median Q3 values are typically between 15 and 20 with interquartile ranges from 10 to 25. Based on these distributions, we selected Q3=5,15,25, and 35 to show Q3 marginal effects at different levels of Q3.

**Figure 2:**
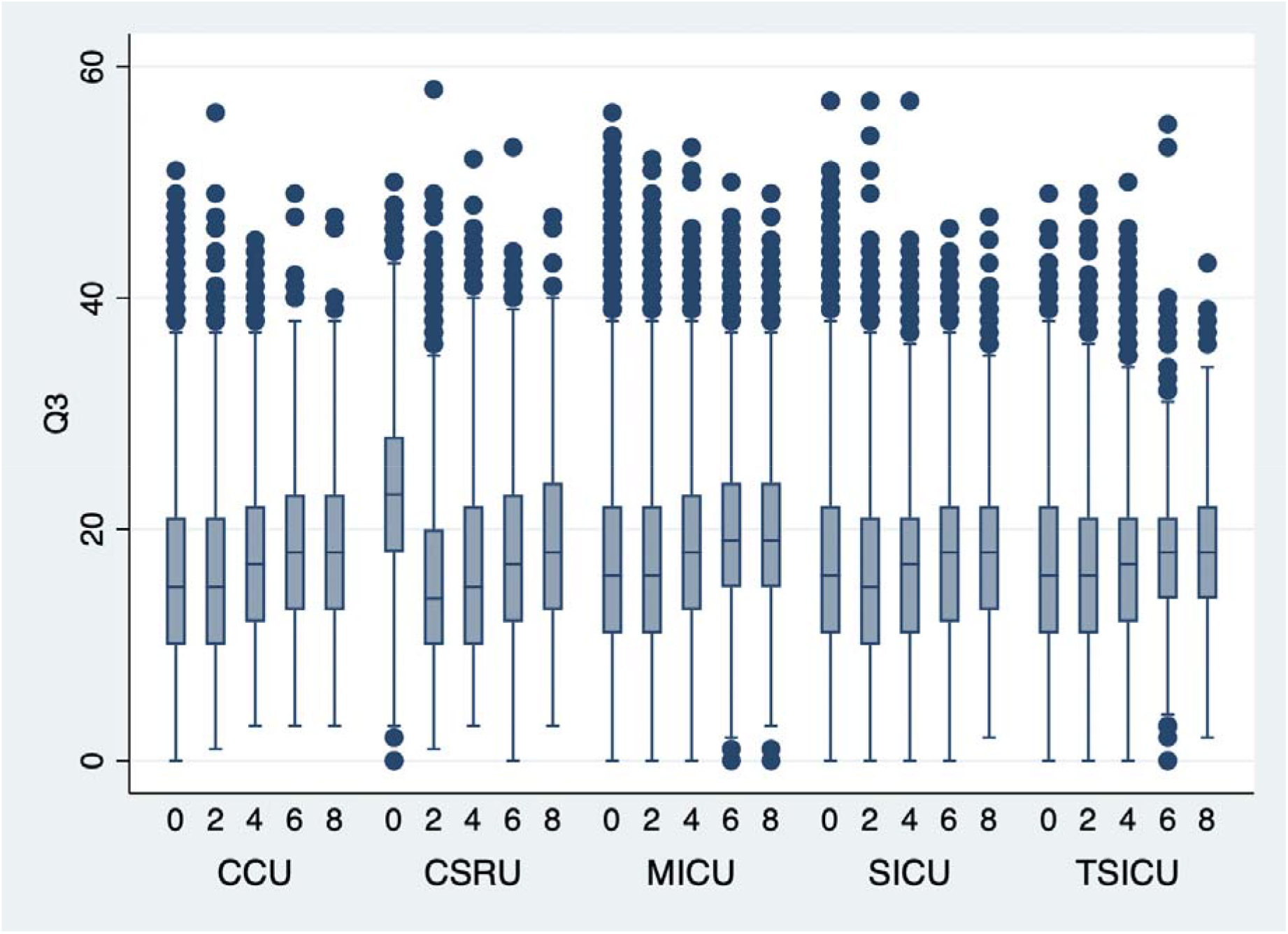
Q3 scores for the first segments of ICU days 0, 2, 4, 6, 8

### ICU mortality

Parameter estimates for Equation B are shown in Table 5. They show that the response is quadratic in Q3 (concave down) through day3, then linear in Q3 from day4 through day9. The marginal effects of Q3 at Q3=5,15, 25, and 35 are included. Pearson-Windmeijer and Stukel tests for miscalibration were insignificant (p>0.05) for all daily models.

**Table 5:** Logistic regression estimates (OR) for ICU mortality (Eq. B)

|  | day0 | day1 | day2 | day3 | day4 | day5 | day6 | day7 | day8 | day9 |
| --- | --- | --- | --- | --- | --- | --- | --- | --- | --- | --- |
| *n* | 28,346 | 20,225 | 13,657 | 9,691 | 7,437 | 5,963 | 4,927 | 4,177 | 3,633 | 3,197 |
| *Q3* | 1.224<sup>‡</sup> | 1.256<sup>‡</sup> | 1.234<sup>‡</sup> | 1.236<sup>‡</sup> | 1.171<sup>‡</sup> | 1.194<sup>‡</sup> | 1.160<sup>‡</sup> | 1.117<sup>‡</sup> | 1.127<sup>‡</sup> | 1.173<sup>‡</sup> |
| *Q3*<sup>2</sup> | .9987<sup>‡</sup> | .9984<sup>‡</sup> | .9989** | .9985** | .9997 | .9992 | .9998 | .9995 | 1.000 | .9993 |
| *Q3 ME at Q3=* |  |  |  |  |  |  |  |  |  |  |
| 5 | 1.21<sup>‡</sup><br>(.012) | 1.24<sup>‡</sup><br>(.015) | 1.22<sup>‡</sup><br>(.017) | 1.22<sup>‡</sup><br>(.020) | 1.17<sup>‡</sup><br>(.021) | 1.18<sup>‡</sup><br>(.026) | 1.16<sup>‡</sup><br>(.028) | 1.16<sup>‡</sup><br>(.031) | 1.13<sup>‡</sup><br>(.030) | 1.17<sup>‡</sup><br>(.032) |
| 15 | 1.18<sup>‡</sup><br>(.006) | 1.20<sup>‡</sup><br>(.008) | 1.19<sup>‡</sup><br>(.009) | 1.18<sup>‡</sup><br>(.010) | 1.16<sup>‡</sup><br>(.010) | 1.17<sup>‡</sup><br>(.013) | 1.15<sup>‡</sup><br>(.014) | 1.15<sup>‡</sup><br>(.015) | 1.14<sup>‡</sup><br>(.015) | 1.15<sup>‡</sup><br>(.016) |
| 25 | 1.15<sup>‡</sup><br>(.004) | 1.16<sup>‡</sup><br>(.004) | 1.17<sup>‡</sup><br>(.005) | 1.15<sup>‡</sup><br>(.006) | 1.15<sup>‡</sup><br>(.006) | 1.15<sup>‡</sup><br>(.007) | 1.15<sup>‡</sup><br>(.008) | 1.14<sup>‡</sup><br>(.008) | 1.14<sup>‡</sup><br>(.008) | 1.14<sup>‡</sup><br>(.008) |
| 35 | 1.12<sup>‡</sup><br>(.008) | 1.12<sup>‡</sup><br>(.009) | 1.14<sup>‡</sup><br>(.011) | 1.12<sup>‡</sup><br>(.013) | 1.14<sup>‡</sup><br>(.015) | 1.13<sup>‡</sup><br>(.017) | 1.15<sup>‡</sup><br>(.019) | 1.13<sup>‡</sup><br>(.020) | 1.15<sup>‡</sup><br>(.021) | 1.12<sup>‡</sup><br>(.020) |
| *MICU*<br>(base) | 1 | 1 | 1 | 1 | 1 | 1 | 1 | 1 | 1 | 1 |
| *SICU* | 0.84** | 0.89 | 0.89 | 0.81* | 0.73** | 0.74** | 0.69** | 0.67<sup>‡</sup> | 0.64<sup>‡</sup> | 0.67** |
| *CCU* | 0.90 | 0.85* | 0.89 | 0.84 | 0.86 | 0.89 | 0.83 | 0.92 | 0.85 | 0.86 |
| *CSRU* | 0.18<sup>‡</sup> | 0.27<sup>‡</sup> | 0.29<sup>‡</sup> | 0.31<sup>‡</sup> | 0.31<sup>‡</sup> | 0.33<sup>‡</sup> | 0.32<sup>‡</sup> | 0.33<sup>‡</sup> | 0.33<sup>‡</sup> | 0.38<sup>‡</sup> |
| *TSICU* | 0.85* | 0.81* | 0.80* | 0.73** | 0.68** | 0.64** | 0.57<sup>‡</sup> | 0.46<sup>‡</sup> | 0.46<sup>‡</sup> | 0.47<sup>‡</sup> |

ME = marginal effect; (SE) = standard error; \* p<0.05, \*\* p<0.01, ‡ p<0.001

Discrimination and calibration tests show that models estimated on Carevue data perform well on Metavision data where they discriminate well (AUROC/c-stat typically > 0.75, Fig 3) and calibrate well (HL p>0.05, Table 6). Spiegelhalter’s tests were insignificant (p>0.05) throughout, further suggesting reasonable discrimination and calibration.

**Figure 3:**
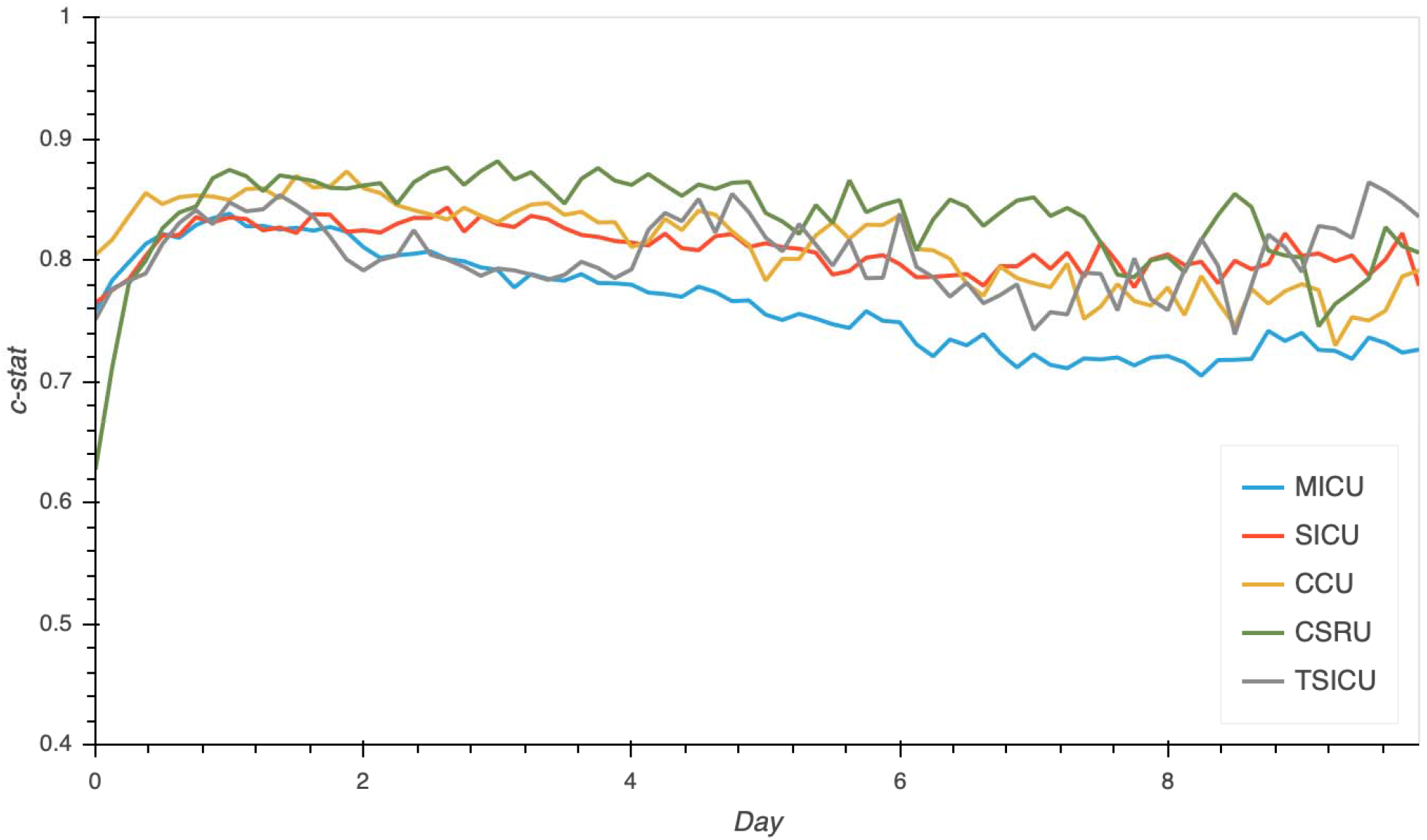
Equation B discrimination testing

**Table 6:**
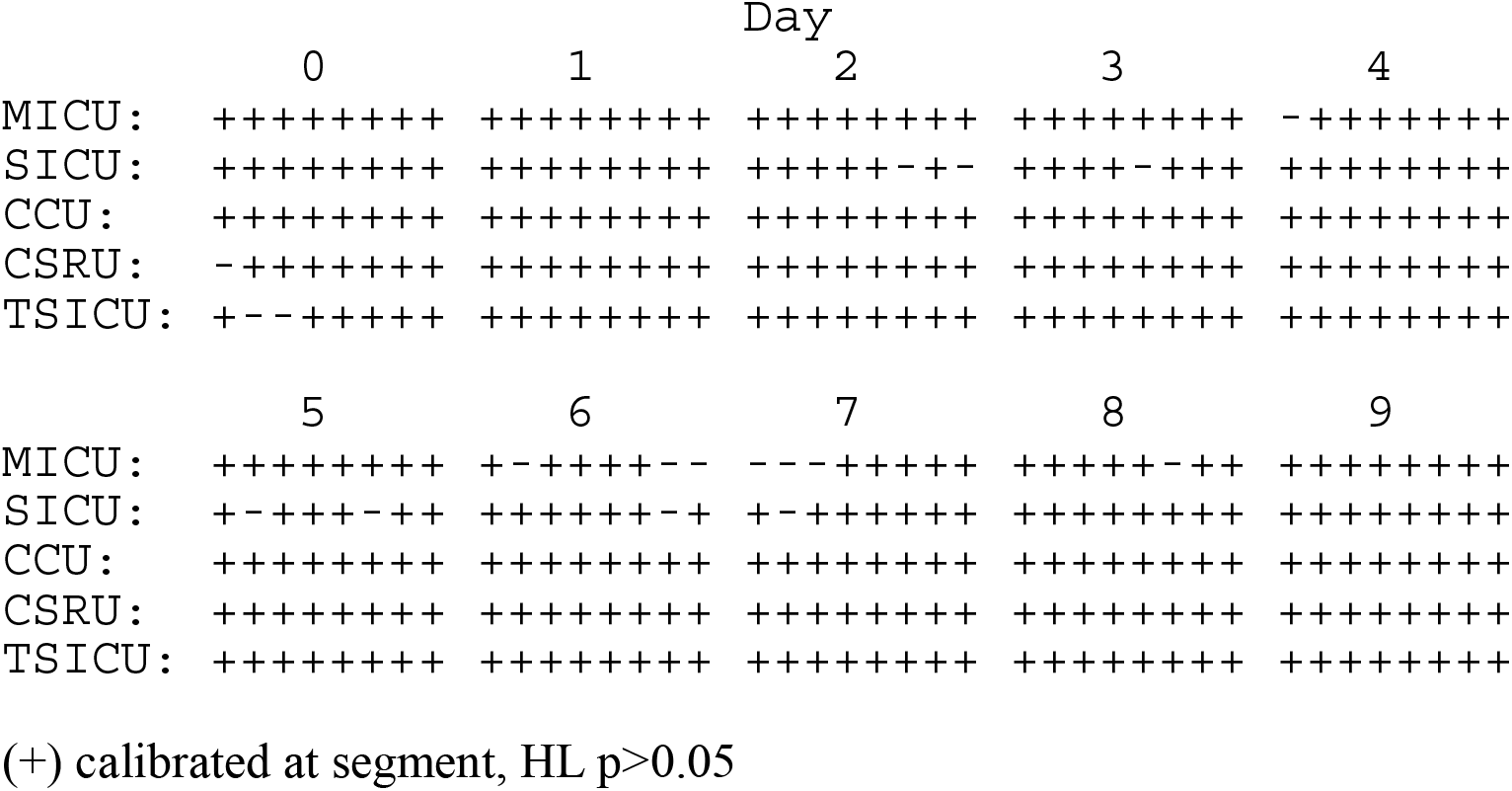
Equation B calibration testing

The average marginal effect (AME) of Q3 on ICU mortality is the average change in probability caused by changing Q3 by one unit while leaving all other covariates unchanged. AME of Q3 by location and day are shown in Table 7. The effects are on the probability scale and are mostly between 1 and 2 percentage points except for the CSRU where AME are smaller.

**Table 7:** Average marginal effects (probability scale) of Q3 on ICU mortality

|  | day0 | day1 | day2 | day3 | day4 | day5 | day6 | day7 | day8 | day9 |
| --- | --- | --- | --- | --- | --- | --- | --- | --- | --- | --- |
| *MICU* | .012<br>(.000) | .013<br>(.000) | .015<br>(.001) | .017<br>(.001) | .018<br>(.001) | .019<br>(.001) | .020<br>(.001) | .020<br>(.001) | .020<br>(.001) | .020<br>(.001) |
| *SICU* | .011<br>(.000) | .012<br>(.000) | .014<br>(.001) | .015<br>(.001) | .015<br>(.001) | .016<br>(.001) | .017<br>(.001) | .016<br>(.001) | .016<br>(.001) | .017<br>(.001) |
| *CCU* | .011<br>(.000) | .012<br>(.001) | .014<br>(.001) | .015<br>(.001) | .017<br>(.001) | .018<br>(.001) | .018<br>(.001) | .019<br>(.001) | .019<br>(.001) | .019<br>(.002) |
| *CSRU* | .003<br>(.000) | .005<br>(.000) | .007<br>(.000) | .008<br>(.001) | .009<br>(.001) | .010<br>(.001) | .010<br>(.001) | .011<br>(.001) | .011<br>(.001) | .012<br>(.001) |
| *TSICU* | .011<br>(.000) | .011<br>(.001) | .013<br>(.001) | .014<br>(.001) | .015<br>(.001) | .015<br>(.001) | .015<br>(.001) | .013<br>(.001) | .013<br>(.001) | .013<br>(.001) |

(SE) = standard error; all estimates significant at $p < 0.001$

### ICU Remaining LOS

The marginal effects of Q3 on rLOS (Eq. C) are shown in Table 8, where for any estimated parameter *B*, the proportional effect of the corresponding variable is exp(*B*)-1. For small |*B*|<0.25, or so, this effect in percentage terms is approximately *B* times 100. For example, the day0 marginal effect of Q3 at Q3=15 is about 5.8% which means that a one-unit increase in Q3 from 15 to 16 increases the remaining ICU LOS by about 5.8%. Location indicators are frequently significant with longer rLOS in the SICU, CSRU, and TSICU than in the MICU and CCU. *Daypart* is sometimes significant, and *weekend* is significant only on day0, where rLOS is about 12% longer than non-weekend segments. The marginal effects of Q3 on rLOS at Q3 levels below 35 are always significant and positive. At Q3=35, the marginal effect is sometimes positive, sometimes negative, and sometimes not significant. Most patients have Q3<25 (Fig 2).

**Table 8:**
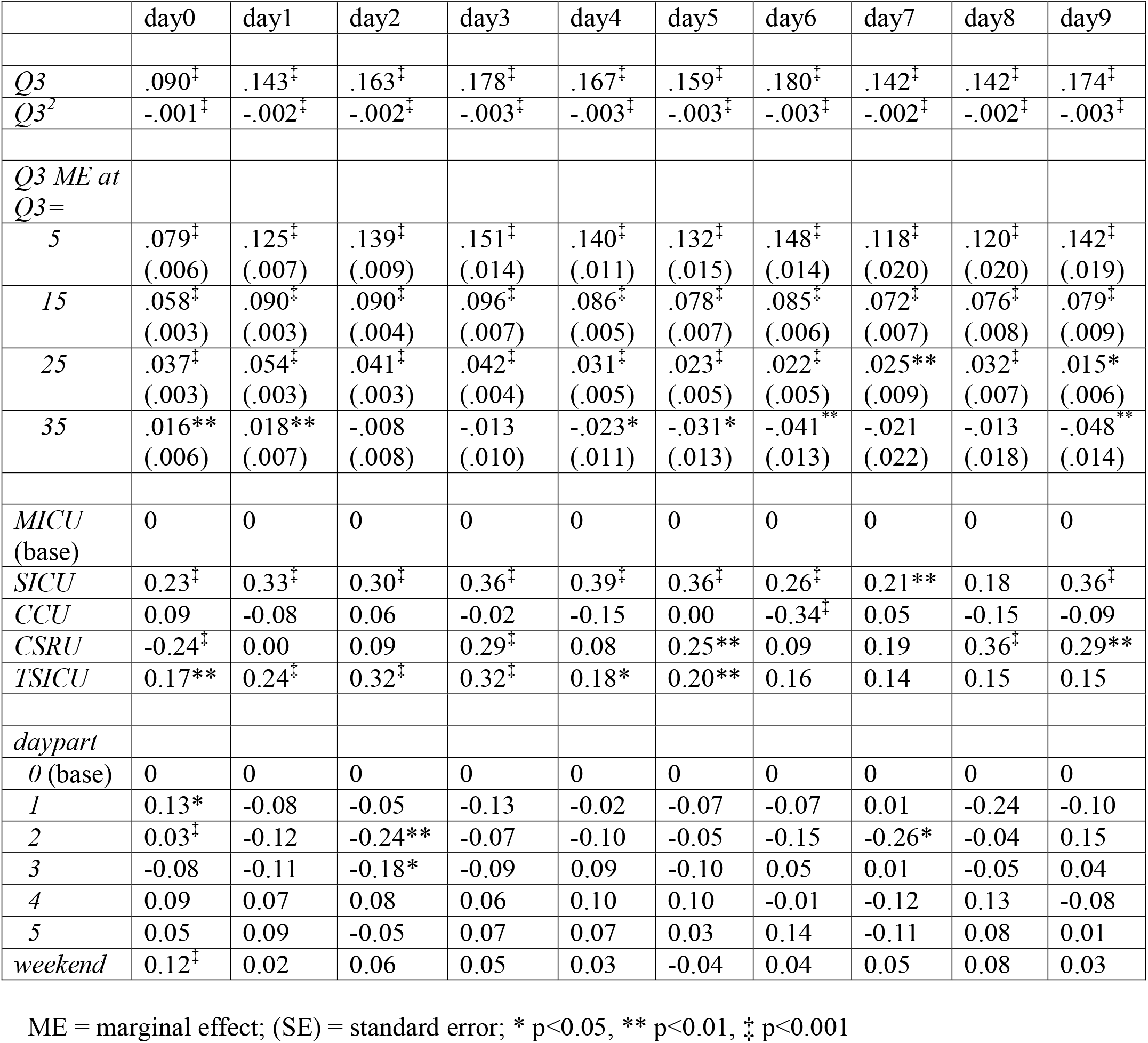
Poisson regression estimates for ICU rLOS (Eq. C)

The average marginal effects of Q3 on rLOS (in hours) are shown in Table 9. They increase over the first few ICU days with most between 5 and 10 hours.

**Table 9:** Average marginal effects (in hours) of Q3 on rLOS

|  | day0 | day1 | day2 | day3 | day4 | day5 | day6 | day7 | day8 | day9 |
| --- | --- | --- | --- | --- | --- | --- | --- | --- | --- | --- |
| *MICU* | 4.3<br>(.19) | 6.2<br>(.24) | 6.5<br>(.30) | 7.9<br>(.42) | 7.5<br>(.42) | 7.5<br>(.51) | 8.7<br>(.63) | 7.8<br>(.82) | 9.6<br>(.87) | 7.4<br>(.75) |
| *SICU* | 5.4<br>(.28) | 8.6<br>(.41) | 8.7<br>(.48) | 11.3<br>(.80) | 11.1<br>(.85) | 10.8<br>(.92) | 11.2<br>(.96) | 9.7<br>(1.1) | 11.5<br>(1.3) | 10.6<br>(1.4) |
| *CCU* | 4.7<br>(.27) | 5.7<br>(.27) | 6.9<br>(.47) | 7.7<br>(.64) | 6.5<br>(.57) | 7.5<br>(.85) | 6.2<br>(.61) | 8.2<br>(1.1) | 8.3<br>(1.4) | 6.8<br>(1.0) |
| *CSRU* | 3.4<br>(.19) | 6.2<br>(.31) | 7.1<br>(.53) | 10.5<br>(.78) | 8.2<br>(.73) | 9.6<br>(1.1) | 9.5<br>(.88) | 9.5<br>(1.4) | 13.8<br>(1.3) | 9.9<br>(1.2) |
| *TSICU* | 5.2<br>(.33) | 7.9<br>(.44) | 8.9<br>(.61) | 10.9<br>(.83) | 9.0<br>(.69) | 9.2<br>(.86) | 10.2<br>(1.1) | 9.0<br>(1.2) | 11.1<br>(1.3) | 8.6<br>(1.1) |

(SE) = standard error; all estimates significant at $p < 0.001$

## Discussion

Acuity scores are used to calculate risk-adjusted mortality rates (actual vs. predicted) for benchmarking and quality improvement [22,23]. Existing ICU acuity scores are typically calculated only at the end of the first ICU day [1-7]. OASIS is non-proprietary and performs about as well as other ICU scores while using fewer variables. However, like other scores, it has only been tested at the end of the first ICU day [10]. Real-time models of ICU acuity use many clinical variables individually instead of one acuity score [8-10], which is limiting if a single score is needed as a dependent variable elsewhere.

In this study, we created Q3, an acuity score computed every 3 hours, and used it in tractable models of ICU mortality and ICU remaining LOS. Q3 is the sum of a new weighted vasopressor score and 8 of the 10 OASIS component scores. Logistic ICU mortality models quadratic in Q3 show no evidence of misspecification and discriminate well (AUROC ∼ 0.72 – 0.85) and calibrate well (Hosemer-Lemeshow p>0.05, Spiegelhalter’s z p>0.05) on newer ICU stays in all ICUs over the first 10 days of the ICU stay. The logistic ICU mortality model is quadratic in Q3 with the quadratic effect different than OR=1 in the first 4 ICU days (Table 5). A one-unit increase in Q3 from a typical level of Q3=15 (Fig 2) affects the odds of ICU mortality by a factor of 1.14 to 1.20 (p<0.001, Table 5) and the ICU remaining LOS by 5.8 to 9.6% (p<0.001, Table 8), depending on the ICU day. The average effect of a one-unit change in Q3 (AME) on ICU mortality are often between 1 and 2 percentage points depending on location and ICU day (Table 7), and the AME of Q3 on ICU remaining LOS are usually between 5 and 10 hours depending on location and ICU day (Table 9).

The marginal effects of Q3 on rLOS are positive at Q3=5, 15, and 25, but at Q3=35, they are inconsistent (Table 8). Since Q3 increases the odds of mortality at all levels of Q3 (Table 5), mortality logically truncates rLOS, and increasing Q3 logically extends LOS not ending in death, the effects of Q3 on rLOS work through mortality and non-mortality causal paths, one plausibly increasing rLOS and the other shortening it. When they are roughly comparable, as they appear to be at high levels of Q3, the marginal effects of Q3 are sometimes positive, sometimes negative, and sometimes insignificant. Ad hoc methods to control for mortality in LOS models include restricting the model to only survivors, assigning large rLOS values to stays censored by death, or adding a mortality indicator. Each of these approaches, however, is problematic since it conditions the regression on a *future* mortality event [24]. Therefore, we chose to leave all stays in and interpret the effects plainly.

We think of Q3 as the residual acuity observable through Q3 components, while location indicators in our models account for average location-specific differences in comorbidities, case mix, and acuity unobserved by Q3. Without location indicators, the average unobserved acuity would be entirely contained in the constant terms of the equations, without location-based resolution. Location indicators work to capture differences between unobserved comorbidities between locations by comparing them to the base level location (MICU). This creates better calibrated models.

Although not shown, we conducted experiments of Q3 without the vasopressor score component. In those experiments, daily estimated models of Equation B show evidence of functional misspecification with several significant Stukel tests (p<0.05), and discriminate less well with c-statistics 1-2 points lower, typically, occasionally dropping below 0.7. This makes sense since *vpres*, the vasopressor score, is extremely significant in addition to other OASIS component scores (Table 4). For these reasons, Q3 with the vasopressor score component is a better score than without it.

To our knowledge, there are several other examples of real-time ICU acuity scores [8-10]. Szolovits uses the MIMIC II database and mortality models that depend on dozens of variables including labs and comorbidities [8]. Overall Day 3 AUROC is about 0.85, comparable to our results (Fig 2, day 2). Location-specific results were not given, and selected model components were not weighted and combined into a single acuity score, like OASIS and Q3. Shickel et. al. use 14 variables including select labs and medication use [9]. It achieves AUROC approaching 0.9 by 96 hours. However, like other deep learning models, its trained neural network is not interpretable. So, while it predicts well, the likely complex, non-linear relationships of input variables encoded by the neural network are currently undiscoverable. Johnson et al use the MIMIC III database to evaluate several real-time mortality prediction models [10]. Features extracted from about 40 clinical variables were computed over 4-hour intervals and were used to predict in-hospital mortality using several model types including logistic regression (LR) and gradient boosting decision trees. The gradient boosting model achieved a high AUROC of 0.93 with LR closely behind at 0.90. No data on model discrimination or calibration by ICU location or at different times during the ICU stay is given, and since many variables are used to capture acuity, no estimated marginal effects of a single acuity score can be given.

We believe that our work contributes to the literature on acuity scores and complements the work of the others, especially the OASIS research team. Q3 is a single score comprised of only 9 variables. When used in tractable and well-behaved models of ICU mortality, Q3 significantly affects the odds of mortality in different ICUs across the first 10 days of the ICU stay (Tabe 5). The marginal effects of Q3 change by the level of Q3 and ICU day (Table 5), and the average marginal effects of Q3 on mortality and ICU remaining LOS are statistically significant and clinically meaningful (Table 7).

Future applications of Q3 include its use as a dependent variable in models that explore the effects of other time-changing variables on ICU acuity, and in estimating the aggregate acuity of subsets of patients over time to help stratify ICU patients in-stay, and to assist with ICU workload analysis.

## Limitations

The MIMIC database contains data from one hospital. However, the data are comprised of many years of observations from different ICUs and patient types.

## Conclusion

Q3 has significant effects on ICU mortality and ICU remaining LOS. Depending on location and ICU day, a one-unit increase in Q3 increases the probability of ICU mortality by or 1-2 percentage points (p<0.001) and ICU remaining LOS by 5 to 10 hours (p<0.001). ICU mortality models that use Q3 discriminate well and are calibrated in different ICUs across the first 10 days of the ICU stay. Given its meaningful effects and favorable performance characteristics, we believe that Q3 could be used as a dependent variable in explanatory models to help elucidate mechanisms of changing ICU acuity.

## Data Availability

Project code is available from the authors.

## Acknowledgements

None

